# Both brain network topology and striatal dopamine depletion mediate the effects of autonomic dysfunction on disease burden of Parkinson’s disease

**DOI:** 10.1101/2023.09.21.23295938

**Authors:** Zhichun Chen, Guanglu Li, Liche Zhou, Lina Zhang, Jun Liu

## Abstract

**Background:** Autonomic dysfunction is one of the most common non-motor symptoms in Parkinson’s disease (PD). Whether autonomic dysfunction contributes to disease progression and brain network abnormalities in PD remain largely unknown. The objective of this study is to evaluate how autonomic dysfunction affects clinical features and brain networks of PD patients.

**Methods:** PD patients from Parkinson’s Progression Markers Initiative (PPMI) database were included if they received magnetic resonance imaging. According to the scores of Scale for Outcomes in Parkinson’s Disease-Autonomic (SCOPA-AUT), PD patients were classified into lower quartile group (SCOPA-AUT score rank: 0%~25%), interquartile group (SCOPA-AUT score rank: 26%~75%), and upper quartile group (SCOPA-AUT score rank: 76%~100%) based on their SCOPA-AUT score quartiles to examine how autonomic dysfunction shapes clinical manifestations and brain networks.

**Results:** PD patients in the upper quartile group showed more severe motor and non-motor symptoms, as well as more deficits in dopamine transporter binding compared to lower quartile group. Additionally, they also showed statistically different topological properties in structural and functional network compared to lower quartile group. Both structural and functional network metrics mediated the effects of autonomic dysfunction on clinical symptoms of PD patients. Reduced dopamine transporter binding also contributed to the effects of autonomic dysfunction on disease burden of PD patients.

**Conclusions:** PD patients with more severe autonomic dysfunction exhibit worse disease and impairment of brain network topology. Both network topology and striatal dopamine depletion mediate the effects of autonomic dysfunction on clinical symptoms of PD patients.

## Introduction

Autonomic dysfunction is an essential non-motor feature of Parkinson’s disease (PD).^[1, 2]^ Recently, an increasing number of studies investigated the specific roles of autonomic dysfunction in the prediction and early diagnosis of PD, which has become a cutting-edge field that directs frontier research.^[1–3]^ Autonomic dysfunction in PD patients includes gastrointestinal dysfunction, cardiovascular dysregulation, urinary disturbance, sexual dysfunction, thermoregulatory aberrance, and pupillo-motor and tear abnormalities.^[1–3]^ It has been reported that over 90% of PD patients will develop autonomic symptoms prior to the onset of Parkinsonian motor symptoms.^[1, 2, 4]^ Autonomic dysfunction was found be associated with older age at onset, longer disease duration, poor initial levodopa treatment response, worse motor symptoms and motor complications, as well as more frequent non-motor symptoms.^[5–8]^ In addition, it has been shown that more severe autonomic dysfunction at baseline was associated with future worse olfaction loss and greater overall disease burden.^[9]^ Moreover, the severity of autonomic symptoms was independently associated with impairments in activities of daily living and health-related quality of life.^[6, 10]^ Therefore, the clinical diagnosis and treatment of autonomic dysfunction is extremely important in PD management.^[1, 2]^ Autonomic dysfunction in PD was associated with multiple risk factors, including age, ^[5, 11]^ male sex, ^[8]^ increased body mass index,^[12]^ total Unified Parkinson’s Disease Rating Scale (UPDRS) score,^[5]^ postural instability,^[13]^ rapid eye movement sleep behavior disorder (RBD),^[14]^ hyposmia,^[5, 15]^ depression,^[5]^ and fatigue.^[5]^ Although tremendous studies have focused on autonomic dysfunction in PD in past decades, the neural mechanisms underlying autonomic failure in PD patients has not yet been identified.^[1, 2, 16]^ According to previous literature, both the degenerations of peripheral and central autonomic systems may be associated with the pathological accumulation of α-synuclein-containing inclusions.^[3, 16–18]^

Functional magnetic resonance imaging (fMRI) has been widely used to explore and investigate how the structural and functional changes in brain contributed to the onset and progression of neuropsychiatric diseases, such as PD, Alzheimer’s disease (AD), bipolar disorder, and schizophrenia.^[19–23]^ Recently, fMRI has helped to identify a series of functional and structural abnormalities significantly associated with autonomic dysfunction in PD patients.^[24–26]^ It has been reported that diffusion changes in medulla oblongata were specifically associated with a lower heart rate and respiratory frequency variability in PD patients.^[26]^ With connectome analysis, Ashraf-Ganjouei *et al*. (2018) revealed that autonomic dysfunction was negatively associated with the integrity of genu of corpus callosum and bilateral cingulum, which suggested structural network contributed to autonomic dysfunction in PD patients.^[24]^ Additionally, it has been shown that compared to PD patients with milder autonomic dysfunction, PD patients with more severe autonomic dysfunction exhibited significantly reduced functional connectivity between the hypothalamus and striatum (caudate, putamen) and thalamus, which was significantly correlated with Scale for Outcomes in Parkinson’s Disease-Autonomic (SCOPA-AUT) scores.^[25]^ These breakthroughs actually enhanced our understanding of autonomic dysfunction in PD patients. However, considering the complexity of central autonomic nervous system,^[1]^ the specific brain networks causally associated with autonomic dysfunction remain largely unclear.

As mentioned above, both structural and functional aberrations in fMRI are associated with autonomic dysfunction in PD patients,^[24–26]^ therefore, we hypothesize that PD patients with worse autonomic dysfunction may exhibit differential topological metrics in structural and functional network compared to PD patients with milder autonomic dysfunction. In order to better decode the associations between brain networks and autonomic dysfunction in PD patients, we classified PD patients into three quartile groups according to their severity of autonomic dysfunction measured by SCOPA-AUT: lower quartile group (Q1, SCOPA-AUT score rank: 0%~25%), interquartile group (Q2–3, SCOPA-AUT score rank: 26%~75%), and upper quartile group (Q4, SCOPA-AUT score rank: 76%~100%). In other words, the goal of this study is to examine whether structural and functional topological metrics are significantly different among different quartile groups and explore the associations between brain topological metrics and autonomic dysfunction. Specifically, our objectives include: (i) to compare clinical assessments among 3 quartile groups; (ii) to compare brain network metrics among 3 quartile groups; (iii) to assess the associations between network metrics and clinical assessments, especially SCOPA-AUT scores; (iv) to examine whether differential topological metrics contribute to the effects of autonomic dysfunction on disease burden of PD patients using mediation analysis.

## Methods

### Participants

The data for the study were collected from the Parkinson’s Progression Markers Initiative (PPMI) database,^[27, 28]^ which can be found at www.ppmi-info.org. This PPMI database consisted of over 400 patients with early PD and 200 control participants. They were subjected to consistent evaluations involving imaging, biological sampling, clinical and behavioral assessments to advance the study of biomarkers associated with PD progression. All procedures were approved by the Institutional Review boards of each participating center, and all participants signed informed consents before enrolling in the study. The eligibility requirements for PD patients in this were as follows: (i) Participants were required to be above the age of 30; (ii) Participants were diagnosed with PD according to the MDS Clinical Diagnostic Criteria for PD; (iii) Participants were required to undergo 3D T1-weighted MPRAGE imaging, resting-state fMRI, and diffusion tensor imaging (DTI) at the same period (within 3 months). The PD patients were excluded from this study if they had noticeable abnormalities in their T1-weighted and T2-weighted MRI scans or had genetic mutations linked to familial PD or were part of the genetic PPMI cohort and prodromal cohort. Based on the criteria stated above, a total of 144 participants diagnosed with PD were chosen for the final analysis. The motor symptoms of PD patients were assessed using various measures such as Hoehn and Yahr stages, Total Rigidity scores, Tremor scores, and the MDS Unified Parkinson’s Disease Rating Scale (MDS-UPDRS). The non-motor assessments included Epworth Sleepiness Scale (ESS), REM Sleep Behavior Disorder Screening Questionnaire (RBDSQ), SCOPA-AUT, 15-item Geriatric Depression Scale (GDS), State-Trait Anxiety Inventory (STAI), Benton Judgment of Line Orientation test (BJLOT), Letter Number Sequencing test (LNS), Semantic Fluency Test (SFT), Symbol Digit Modalities Test (SDMT), and Montreal Cognitive Assessment (MoCA). The patients also received [123I] FP-CIT SPECT scans, which were evaluated following the guidelines outlined in the technical manual of the PPMI study (http://ppmi-info.org/). The SBRs for both sides of caudate, putamen, and striatum were calculated based on SPECT scans. The calculation of the SBRs was performed using the formula: (target region/reference region)-1 in which occipital lobe served as the reference region. In order to investigate the effects of autonomic dysfunction on clinical features and brain networks, PD patients (n = 144) were divided into lower quartile group (Q1, SCOPA-AUT rank: 0%~25%, SCOPA-AUT range: 0~6), interquartile group (Q2-3, SCOPA-AUT rank: 26%~75%, SCOPA-AUT range: 6~13), and upper quartile group (Q4, SCOPA-AUT rank: 76%~100%, SCOPA-AUT range: 14~27) based on their SCOPA-AUT quartiles. The clinical characteristics of participants in 3 groups were displayed in Table 1 and Figure 1. The similar method for subgroup classifications according to the quartiles of continuous variables have been documented previously.^[29–32]^

**Figure 1:**
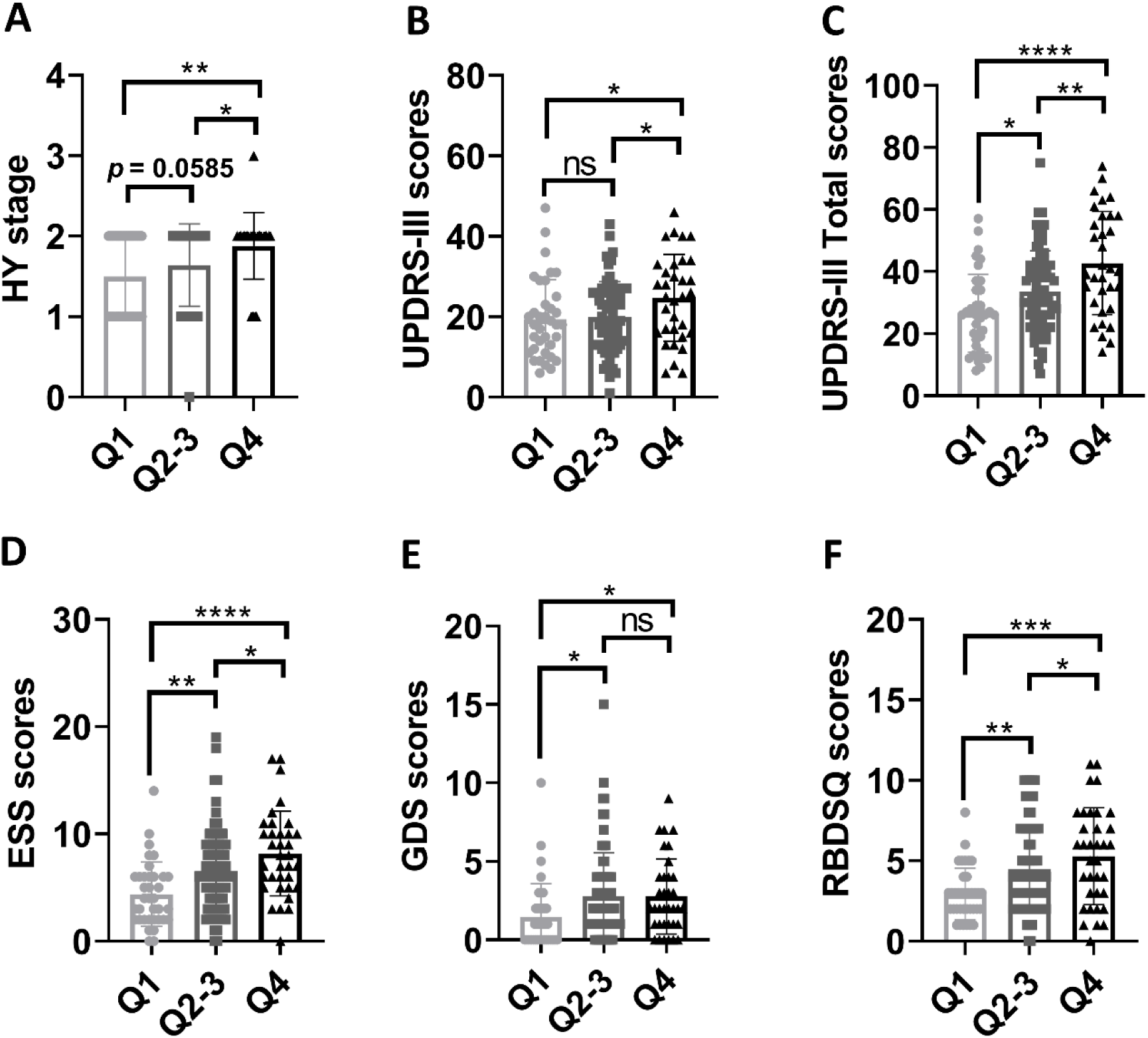
Group differences in clinical assessments. (A-F) Group differences in HY stages (A), UPDRS-III scores (B), total UPDRS scores (C), ESS scores (D), GDS scores (E), and RBDSQ scores (F). One-way ANOVA followed by Tukey’s post hoc test (Q1 group *vs* Q2-3 group *vs* Q4 group) were conducted to compare clinical variables. *p* < 0.05 was considered statistically significant. \**p* < 0.05, \*\**p* < 0.01, \*\*\**p* < 0.001, \*\*\*\**p* < 0.0001. Abbreviations: HY, Hoehn & Yahr stage; UPDRS, Unified Parkinson’s Disease Rating Scale; ESS, Epworth Sleepiness Scale; GDS, 15-item Geriatric Depression Scale; RBDSQ, REM Sleep Behavior Disorder Screening Questionnaire.

**Table 1:**
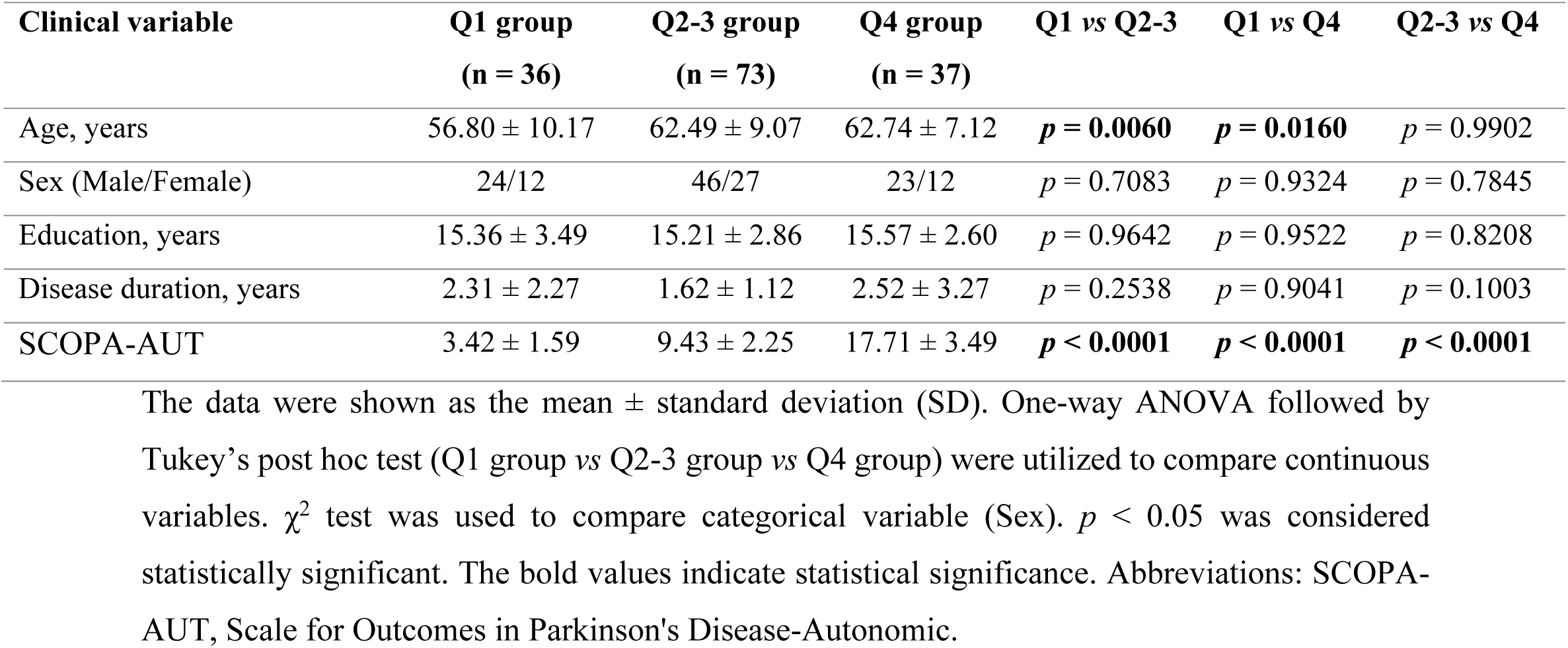
The demographic and clinical data for each quartile group.

### Image acquisition

The imaging data were obtained using Siemens 3T Trio or Verio scanners from Siemens Medical Solutions in Malvern, PA. The rs-fMRI was acquired using echo-planar imaging sequence (Repetition time [TR] 2,400 ms, Echo time (ET) 25 ms, 40 slices with a thickness of 3.25 mm, Flip angle 80.0°, and Field of view 222 × 222 mm). The T1-weighted MRI images in three dimensions were obtained utilizing a magnetization-prepared rapid acquisition gradient echo sequence (TR 2300 ms, TE 2.98 ms, Voxel size 1 mm^3^, Slice thickness 1.2 mm, twofold acceleration, and sagittal-oblique angulation). The DTI was acquired with the following parameters: TR = 8,400-8,800 ms, TE = 88 ms, Voxel size = 2 mm^3^, Slice thickness = 2 mm, 64 directions, b-value = 1000 s/mm^2^.

### Imaging preprocessing

The FMRIB Software Library toolkit (FSL, https://fsl.fmrib.ox.ac.uk/fsl/fslwiki) was utilized to preprocess DTI images from 144 PD patients. In brief, the DTI images were initially corrected for head motions, eddy-current distortions, and susceptibility artifacts. Afterwards, DTI measurements including fractional anisotropy (FA), mean diffusivity (MD), axial diffusivity, and radial diffusivity, were derived. Finally, the individual processed images were additionally reconstructed in the standard MNI space for comparisons among participants.

The resting-state fMRI images were preprocessed using SPM12 (http://www.fil.ion.ucl.ac.uk/spm/software/spm12/) and GRETNA toolbox (https://www.nitrc.org/projects/gretna/). The preprocessing of rs-fMRI images went through several steps, including slice-time correction; realignment and unwarping; segmentation of gray matter, white matter, and CSF; spatial normalization to the Montreal Neurological Institute template; and smoothing based on a 4-mm Gaussian kernel. Next, nuisance variable regression was conducted, where the first 5 principal components derived from the segmented white matter and CSF, the 6 motion realignment parameters, along with their derivatives and outlier volumes were removed from the signal through regression. Next, the data underwent bandpass filtering on the residual signals within the frequency range of 0.01–0.1 Hz. According to prior studies,^[33–36]^ we removed participants from our sample who had a frame-wise displacement (FD) of over 0.5 mm and head rotation of more than 2° caused by head motions. There were no significant differences in FD values among the 3 quartile groups (*p* > 0.05).

### Network construction

An open and free MATLAB toolkit, PANDA (http://www.nitrc.org/projects/panda/), was utilized to perform deterministic fiber tractography to generate the structural network. Briefly, Fiber Assignment by Continuous Tracking (FACT) algorithm was used to construct white matter fibers connecting all 90 cortical and subcortical nodes in the Automated Anatomical Labeling (AAL) atlas. The cut-off for FA skeleton was established as 0.20, and a 45° threshold was applied for fiber angle. After conducting white matter tractography, a white matter network matrix was created for each participant using fiber number (FN) as edges.

The generation of functional network matrix was performed as follows. At first, the functional connectivity for each pair of nodes in AAL atlas was calculated as the Pearson correlation coefficient of their time series data. Afterwards, Fisher’s r-to-z conversions for the correlation coefficients were performed to improve the normal distribution of functional connectivity. In the end, a 90 x 90 functional matrix was constructed for each PD participant.

### Graph-based network analysis

The topological network metrics of structural and functional networks were computed using GRETNA (https://www.nitrc.org/projects/gretna/). A range of network sparsity thresholds (0.05 ~ 0.50 with a step size of 0.05) was used to calculate the global and nodal network properties. The global network metrics included: global efficiency, local efficiency, and small-worldness properties. The nodal network metrics included: nodal betweenness centrality, nodal degree centrality, nodal clustering coefficient (Cp), nodal efficiency, nodal local efficiency, and nodal shortest path length. The mathematical definitions for all network properties have been provided by previous studies.^[37–39]^

### Statistical analysis

#### Comparison of clinical variables

One-way ANOVA followed by Tukey’s post hoc test (Q1 group *vs* Q2-3 group *vs* Q4 group) was utilized to identify group differences of continuous variables. χ^2^ test was used to analyze categorical variables. *p* < 0.05 was considered statistically significant.

#### Comparison of global network strength

Network-Based Statistic (NBS, https://www.nitrc.org/projects/nbs/) toolbox was utilized to compare the global network strength of brain networks among 3 quartile groups. *p* < 0.05 after false discovery rate (FDR) correction was considered statistically significant. Age, sex, years of education, and disease duration, were included as covariates during NBS analysis.

#### Comparison of network metrics

Two-way ANOVA test followed by FDR corrections was utilized to compared global and nodal network metrics. *p* < 0.05 after FDR correction was considered statistically significant.

### Association analysis

The univariant correlations between SCOPA-AUT scores and clinical variables or network metrics were performed with Pearson correlation method. The multivariate regression analysis to examine the connections between SCOPA-AUT scores and clinical variables or network metrics, taking into account age, sex, disease duration, and education level. For associations between SCOPA-AUT scores and clinical variables, *p* < 0.05 was considered statistically significant. For relationships between SCOPA-AUT scores and graphical network metrics, *p* < 0.05 after FDR correction was considered statistically significant.

### Mediation analysis

IBM SPSS Statistics Version 26 was utilized to perform the mediation analysis. The independent variables during mediation analysis included SCOPA-AUT scores. The dependent variables included scores of clinical assessments (HY, UPDRS-III, Total UPDRS, SDMT and MoCA). The mediators were topological network metrics and SBRs of bilateral caudate, putamen, or striatum. We modeled the indirect effects of network metrics or striatum SBRs on the relationships between SCOPA-AUT scores (autonomic dysfunction) and clinical assessments. Age, sex, disease duration, and years of education were enrolled as covariates during the mediation analysis. *p* < 0.05 was considered statistically significant.

## Results

### Group differences of clinical variables

As shown in Table 1, compared to patients in Q1 group (n = 36), Q2-3 (n = 73) and Q4 group (n = 37) showed higher age (*p* = 0.0060 and *p* = 0.0160, respectively) and SCOPA-AUT scores (both *p* < 0.0001), whereas sex, years of education, and disease duration were not statistically different among 3 quartiles. In addition, Figure 1 showed Q4 group exhibited higher HY stages (*p* < 0.01), UPDRS-III scores (*p* < 0.05), total UPDRS scores (*p* < 0.0001), ESS scores (*p* < 0.0001), GDS scores (*p* < 0.05), and RBDSQ scores (*p* < 0.001) compared to Q1 group. Q4 group also showed higher HY stages (*p* < 0.05), UPDRS-III scores (*p* < 0.05), total UPDRS scores (*p* < 0.01), ESS scores (*p* < 0.05), and RBDSQ scores (*p* < 0.05) compared to Q2-3 group. Compared to Q1 group, Q2-3 group showed higher HY stages (*p =* 0.0585), total UPDRS scores (*p* < 0.05), ESS scores (*p* < 0.01), GDS scores (*p* < 0.05), and RBDSQ scores (*p* < 0.01). Furthermore, Q2-3 and Q4 group exhibited much lower SBRs in right caudate, left caudate, right putamen, left putamen, right striatum, left striatum, bilateral caudate, bilateral putamen, and bilateral striatum compared to Q1 group (Figure S1).

### Associations between SCOPA-AUT scores and clinical variables

For demographic variables, age was significantly associated with SCOPA-AUT scores (β = 0.14, *p* = 0.0060) while sex, years of education, and disease duration were not significantly associated with SCOPA-AUT scores. For clinical assessments, as shown in Table 2, SCOPA-AUT scores were positively associated with HY stages (*p* < 0.01), UPDRS-III scores (*p* < 0.05), total UPDRS scores (*p* < 0.0001), ESS scores (*p* < 0.0001), GDS scores (*p* < 0.01), RBDSQ scores (*p* < 0.0001), STAI scores (*p* < 0.001), and negatively associated with SDMT (*p* < 0.05) and MoCA scores (*p* < 0.01). In addition, SCOPA-AUT scores were negatively associated with SBRs in right caudate, left caudate, right putamen, left putamen, right striatum, left striatum, bilateral caudate, bilateral putamen, and bilateral striatum (Table 2).

**Table 2:** Associations between SCOPA-AUT scores and clinical assessments.

| Clinical assessments |  |  |  |  |  |  |  |  |  |
| --- | --- | --- | --- | --- | --- | --- | --- | --- | --- |
| Variables<br>Statistics | HY stage | Tremor | Rigidity | UPDRS-III | UPDRS-Total | ESS | GDS | RBDSQ |  |
| $\beta$ value | $\beta = 0.02$ | $\beta = -0.01$ | $\beta = 0.07$ | $\beta = 0.37$ | $\beta = 1.11$ | $\beta = 0.29$ | $\beta = 0.12$ | $\beta = 0.19$ | |
| $p$-value | $p = 0.0060$ | $p = 0.7964$ | $p = 0.1065$ | $p = 0.0136$ | $p < 0.0001$ | $p < 0.0001$ | $p = 0.0032$ | $p < 0.0001$ | |
| Clinical assessments |  |  |  |  |  |  |  |  |  |
| Variables<br>Statistics | STAI | BJLOT | SFT | SDMT | LNS | MoCA | HVLT-R<br>Immediate Recall | HVLT-R<br>Delayed Recall |  |
| $\beta$ value | $\beta = 1.06$ | $\beta = -0.03$ | $\beta = -0.22$ | $\beta = -0.32$ | $\beta = -0.06$ | $\beta = -0.10$ | $\beta = -0.06$ | $\beta = -0.03$ | |
| $p$-value | $p = 0.0003$ | $p = 0.2897$ | $p = 0.1906$ | $p = 0.0343$ | $p = 0.1134$ | $p = 0.0089$ | $p = 0.4715$ | $p = 0.5207$ | |
| Clinical assessments |  |  |  |  |  |  |  |  |  |
| SBRs<br>Statistics | Right caudate | Left caudate | Right putamen | Left putamen | Right striatum | Left striatum | Bilateral<br>caudate | Bilateral<br>putamen | Bilateral<br>striatum |
| $\beta$ value | $\beta = -0.03$ | $\beta = -0.03$ | $\beta = -0.01$ | $\beta = -0.01$ | $\beta = -0.04$ | $\beta = -0.05$ | $\beta = -0.03$ | $\beta = -0.01$ | $\beta = -0.02$ |
| $p$-value | $p < 0.0001$ | $p = 0.0002$ | $p = 0.0342$ | $p = 0.0049$ | $p = 0.0005$ | $p = 0.0003$ | $p < 0.0001$ | $p = 0.0038$ | $p < 0.0001$ |

The data were shown as the $\beta$ and $p$ -values derived from multivariate regression analysis with age, sex, years of education, and disease duration as covariates. The motor function examination was assessed in ON state. $p < 0.05$ was considered statistically significant. The bold values indicate statistical significance. Abbreviations: HY, Hoehn & Yahr; UPDRS, Unified Parkinson's Disease Rating Scale; ESS, Epworth Sleepiness Scale; RBDSQ, REM Sleep Behavior Disorder Screening Questionnaire; GDS, Geriatric Depression Scale; SCOPA-AUT, Scale for Outcomes in Parkinson's Disease-Autonomic; SDMT, Symbol Digit Modalities Test; LNS, Letter Number Sequencing; SFT, Semantic Fluency Test Score; STAI, State-Trait Anxiety Inventory; BJLOT, Benton Judgement of Line Orientation; MoCA, Montreal Cognitive Assessment; HVLT-R, Hopkins Verbal Learning Test-Revised; SBR, Striatal binding ratio.

### Group differences in network metrics

We found global network metrics in structural network and functional network were not statistically different among 3 quartiles (all *p* > 0.05). For structural nodal network metrics, compared to Q1 group, Q4 group showed lower betweenness centrality (FDR-corrected *p* < 0.05 in right superior frontal gyrus and left calcarine; Figure 2A) and degree centrality (FDR-corrected *p* < 0.05 in left superior frontal gyrus, right superior frontal gyrus, and left calcarine; Figure 2B). Compared to Q1 and Q2-3 group, Q4 group displayed higher nodal Cp (FDR-corrected *p* < 0.05 in left middle orbitofrontal gyrus, left Heschl, and right Heschl; Figure 2C) and nodal local efficiency (FDR-corrected *p* < 0.05 in left middle orbitofrontal gyrus, left Heschl, and right Heschl; Figure 2C). Compared to Q1 group, Q2-3 group and Q4 group exhibited much lower nodal efficiency (FDR-corrected *p* < 0.05 in left superior frontal gyrus, right superior frontal gyrus, right superior parietal gyrus; Figure 2D) and grater nodal shortest path length (FDR-corrected *p* < 0.05 in left middle orbitofrontal gyrus and bilateral superior medial frontal gyri; Figure 2E).

**Figure 2:**
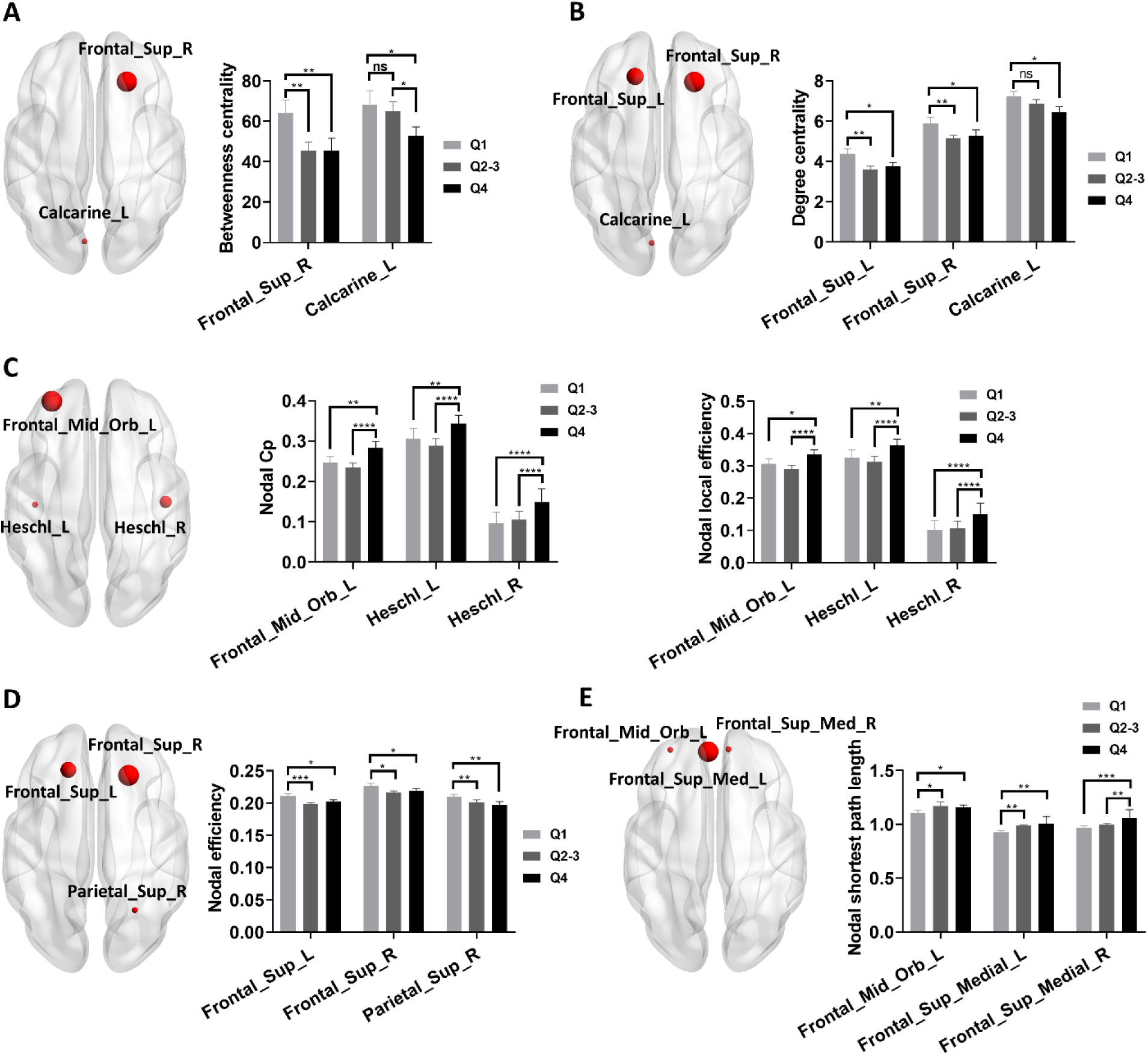
Group differences in nodal network metrics of structural network. (A-E) Group differences of nodal betweenness centrality (A), nodal degree centrality (B), nodal Cp and nodal local efficiency (C), nodal efficiency (D), and nodal shortest path length (E). Two-way ANOVA test with FDR corrections was performed. FDR-corrected *p* < 0.05 was considered statistically significant. \**p* < 0.05, \*\**p* < 0.01, \*\*\**p* < 0.001, \*\*\*\**p* < 0.0001. Abbreviations: Cp, Clustering coefficient.

For functional network metrics, compared to Q1 group, Q4 group exhibited higher degree centrality (FDR-corrected *p* < 0.05 in right middle orbitofrontal gyrus, right posterior cingulate cortex, and left posterior cingulate cortex; Figure 3A), nodal efficiency (FDR-corrected *p* < 0.05 in right posterior cingulate cortex and left posterior cingulate cortex; Figure 3B), nodal local efficiency (FDR-corrected *p* < 0.05 in right rectus, left rectus, left posterior cingulate cortex; Figure 3C), and lower nodal shortest path length (FDR-corrected *p* < 0.05 in left anterior cingulate cortex, left posterior cingulate cortex, right posterior cingulate cortex; Figure 3A).

**Figure 3:**
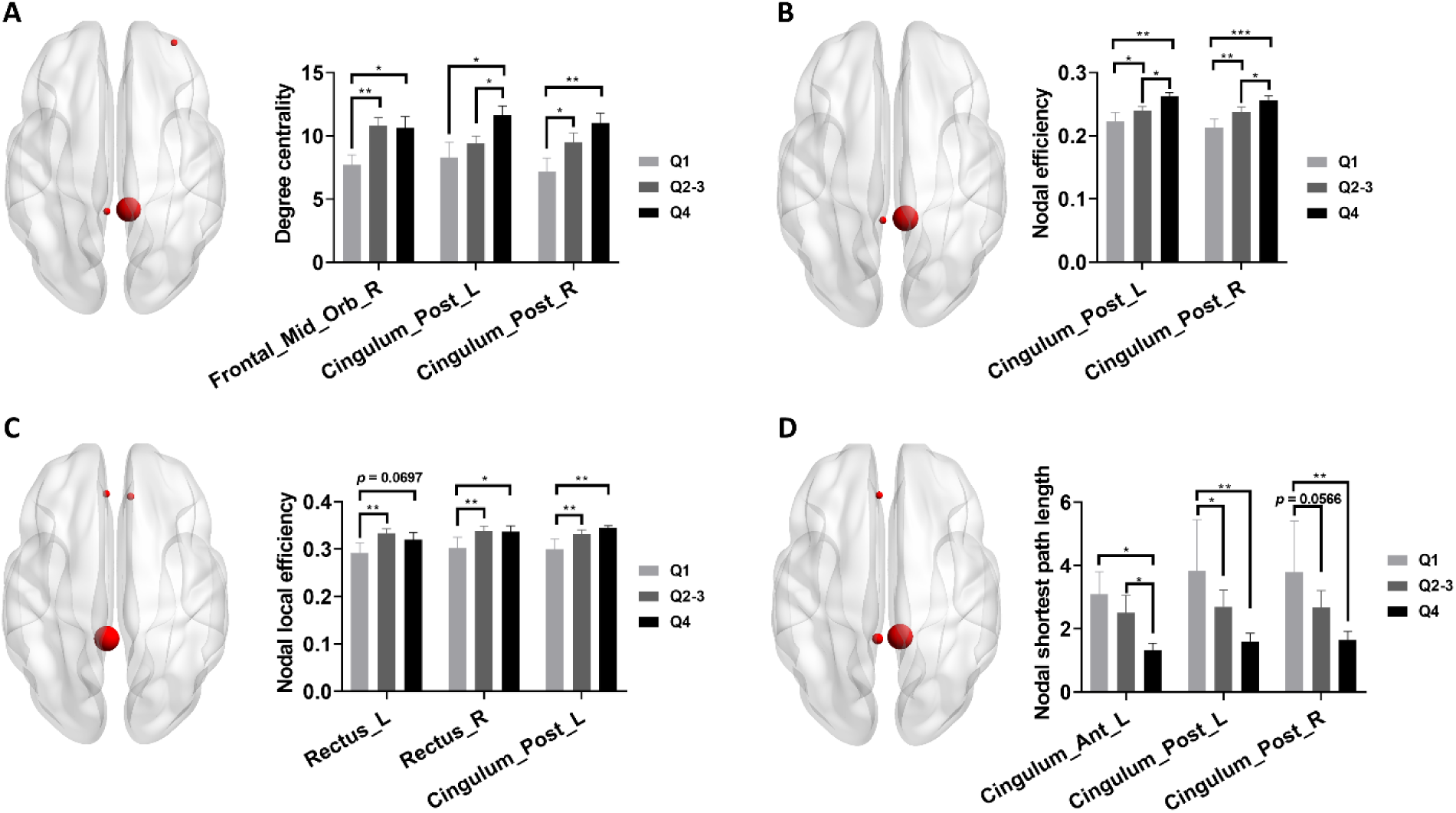
Group differences in nodal network metrics of functional network. (A-D) Group differences of nodal degree centrality (A), nodal efficiency (B), nodal local efficiency (C), and nodal shortest path length (D). Two-way ANOVA test with FDR corrections was performed. FDR-corrected *p* < 0.05 was considered statistically significant. \**p* < 0.05, \*\**p* < 0.01, \*\*\**p* < 0.001.

### Associations between SCOPA-AUT scores and nodal network metrics

For structural network metrics, we found degree centrality in left calcarine was negatively associated with SCOPA-AUT scores, which was independent of age, sex, years of education, and disease duration (Figure S2A). For functional network metrics, we found degree centrality in left posterior cingulate cortex (Figure S2B) and right posterior cingulate cortex (Figure S2C) were positively associated with SCOPA-AUT scores. Additionally, nodal efficiency in left posterior cingulate cortex (Figure S2C) and right posterior cingulate cortex (Figure S2D) were also positively associated with SCOPA-AUT scores.

### Associations between nodal network metrics and clinical assessments

Among above differential network metrics shown in Figure 2 and Figure 3, we found degree centrality in left calcarine was negatively associated with HY stages (Figure 4A), UPDRS-III scores (Figure 4B), total UPDRS scores (Figure 4C), ESS scores (Figure 4D) and positively associated with SDMT (Figure 4E) and MoCA scores (Figure 4F).

**Figure 4:**
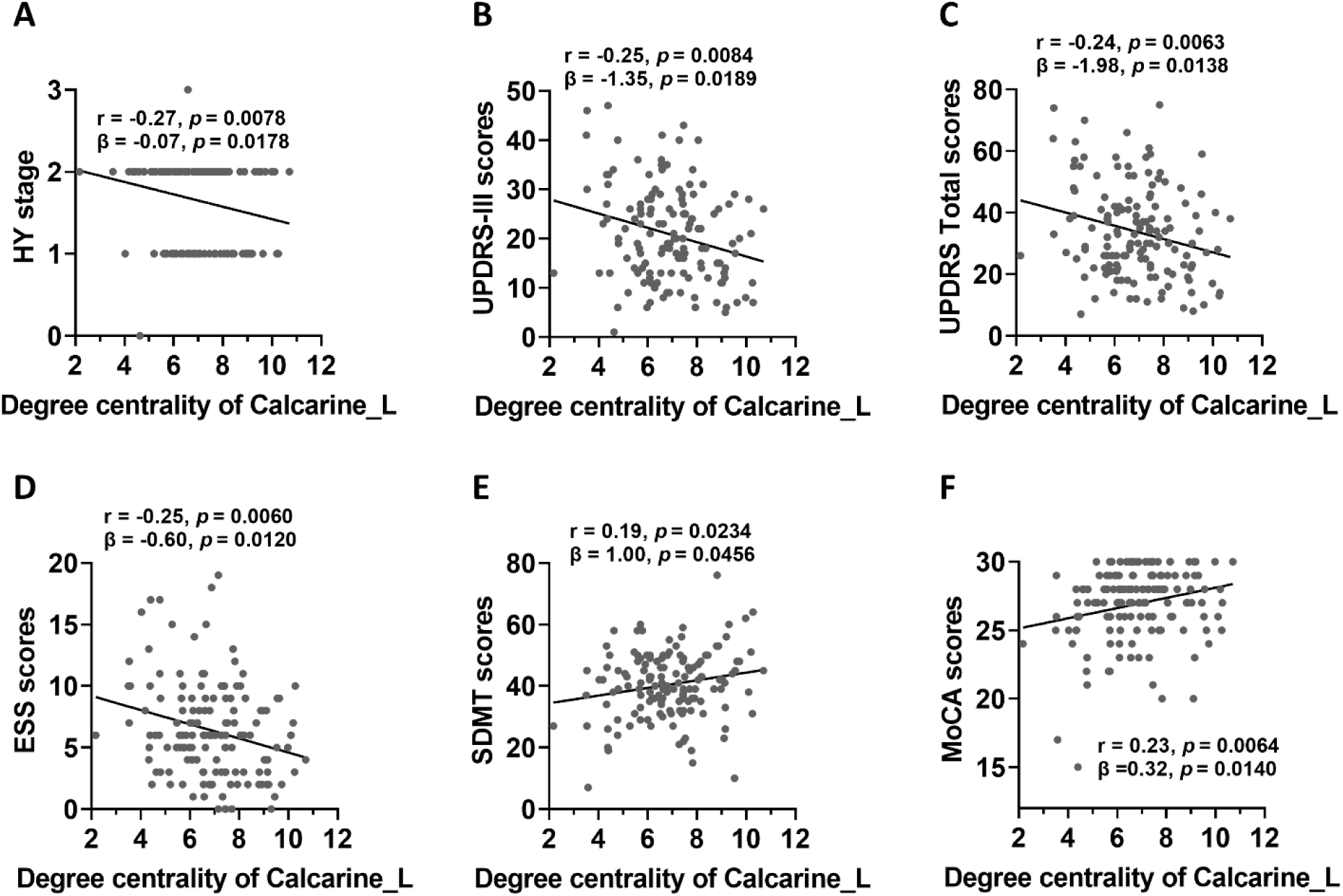
Associations between nodal network metrics and clinical assessments. (A-D) Degree centrality of left calcarine was negatively associated with HY stages, UPDRS-III scores, total UPDRS scores, and ESS scores (FDR-corrected *p* < 0.05 in both Pearson correlation analysis and multivariate regression analysis). (E-F) Degree centrality of left calcarine was positively associated with SDMT scores and MoCA scores (FDR-corrected *p* < 0.05 in both Pearson correlation analysis and multivariate regression analysis). The association analysis between graphical network metrics and clinical assessments was conducted by Pearson correlation method and multivariate regression analysis with age, sex, disease duration, and years of education as covariates. FDR-corrected *p* < 0.05 was considered statistically significant. Abbreviations: HY, Hoehn & Yahr stage; UPDRS, Unified Parkinson’s Disease Rating Scale; ESS, Epworth Sleepiness Scale; SDMT, Symbol Digit Modalities Test; MoCA, Montreal Cognitive Assessment.

### Mediation analysis

Because degree centrality in left calcarine of structural network was associated with both motor and non-motor symptoms of PD patients, we examined whether degree centrality in left calcarine mediated the associations between autonomic dysfunction (measured by SCOPA-AUT scores) and clinical features. As shown in Figure 5, degree centrality in left calcarine mediated the positive association between autonomic dysfunction and HY stages (Figure 5A) and UPDRS-III scores (Figure 5B). In addition, degree centrality in left calcarine also mediated the negative association between autonomic dysfunction and MoCA scores (Figure 5C). For functional network metrics, we found degree centrality and nodal efficiency in bilateral posterior cingulate cortices mediated the negative association between autonomic dysfunction and SDMT scores (Figure 6).

**Figure 5:**
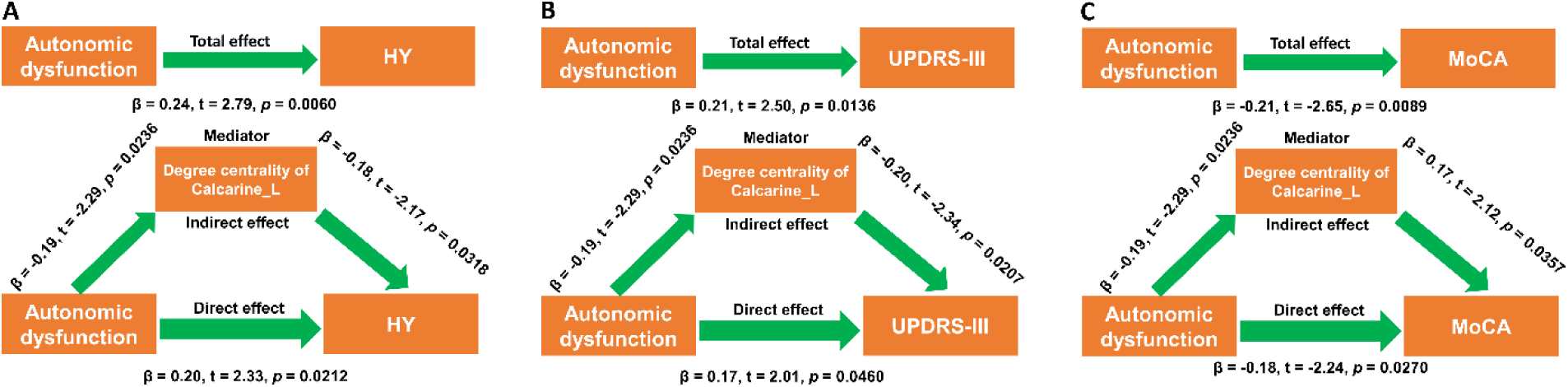
Degree centrality of left calcarine in structural network mediated the effects of autonomic dysfunction on motor and non-motor symptoms. (A-C) Degree centrality of left calcarine mediated the effects of autonomic dysfunction on HY stage (A), UPDRS-III scores (B), and MoCA scores (C). During the mediation analysis, age, sex, disease duration, and years of education were included as covariates. *p* < 0.05 was considered statistically significant. Abbreviations: HY, Hoehn & Yahr stage; UPDRS, Unified Parkinson’s Disease Rating Scale; MoCA, Montreal Cognitive Assessment.

**Figure 6:**
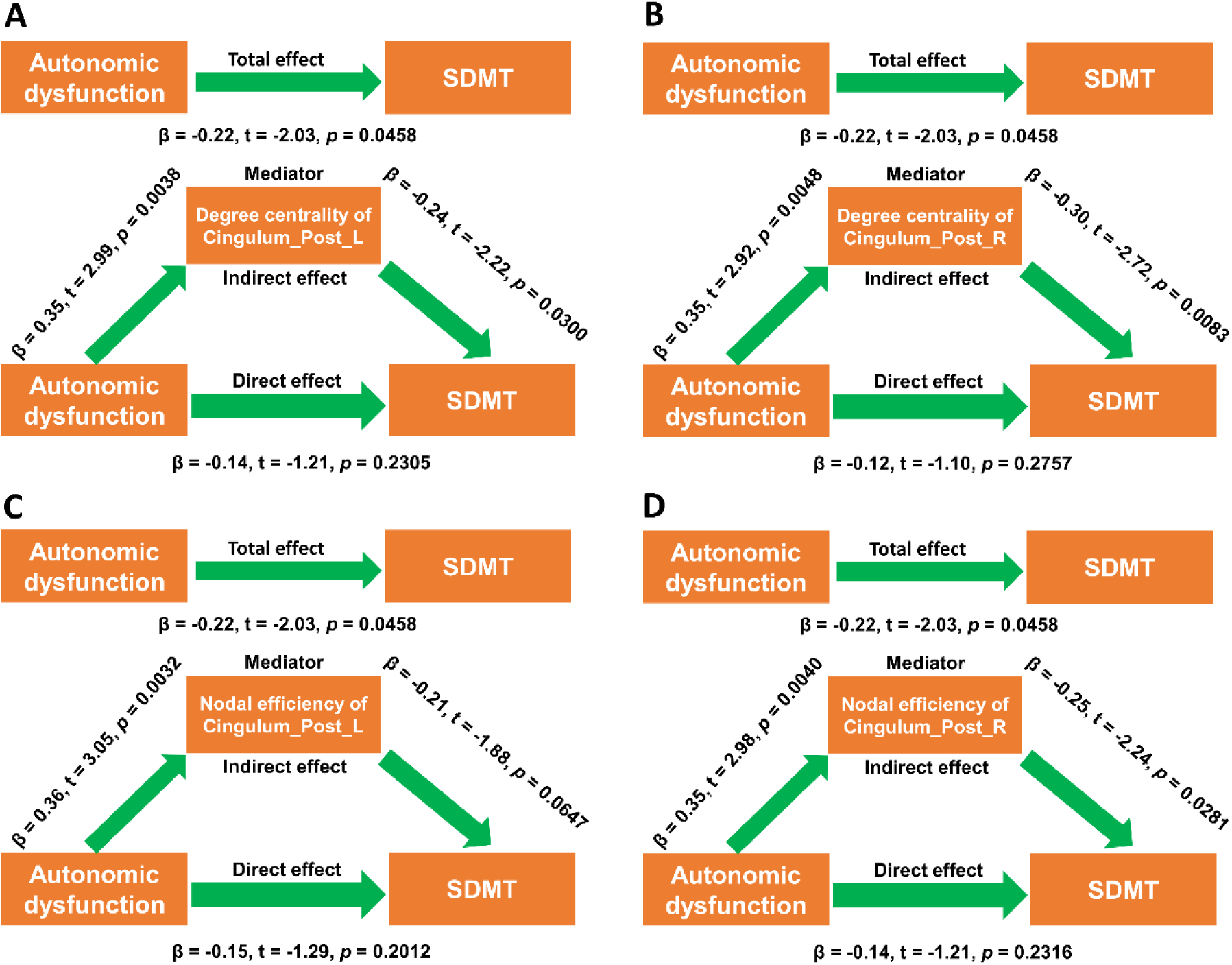
Degree centrality and nodal efficiency of bilateral posterior cingulate cortex in functional network mediated the effects of autonomic dysfunction on SDMT scores. (A-B) Degree centrality of left posterior cingulate cortex (A) and right posterior cingulate cortex (B) mediated the effects of autonomic dysfunction on SDMT scores. (C-D) Nodal efficiency of left posterior cingulate cortex (C) and right posterior cingulate cortex (D) mediated the effects of autonomic dysfunction on SDMT scores. During the mediation analysis, age, sex, disease duration, and years of education were included as covariates. *p* < 0.05 was considered statistically significant. Abbreviations: SDMT, Symbol Digit Modalities Test.

Because SCOPA-AUT scores were negatively associated with striatum SBRs, we examined whether striatum SBRs also mediated the effects of autonomic dysfunction on clinical symptoms. As shown in Figure 7, we found SBR of right putamen specifically mediated the effects of autonomic dysfunction on UPDRS-III scores (Figure 7A) while other striatal regions failed to reach statistical significance. Additionally, SBRs in right caudate (Figure 7B), right putamen (Figure 7C), right striatum (Figure 7D), bilateral putamen (Figure 7E), and bilateral striatum (Figure 7F) mediated the effect of autonomic dysfunction on total UPDRS scores.

**Figure 7:**
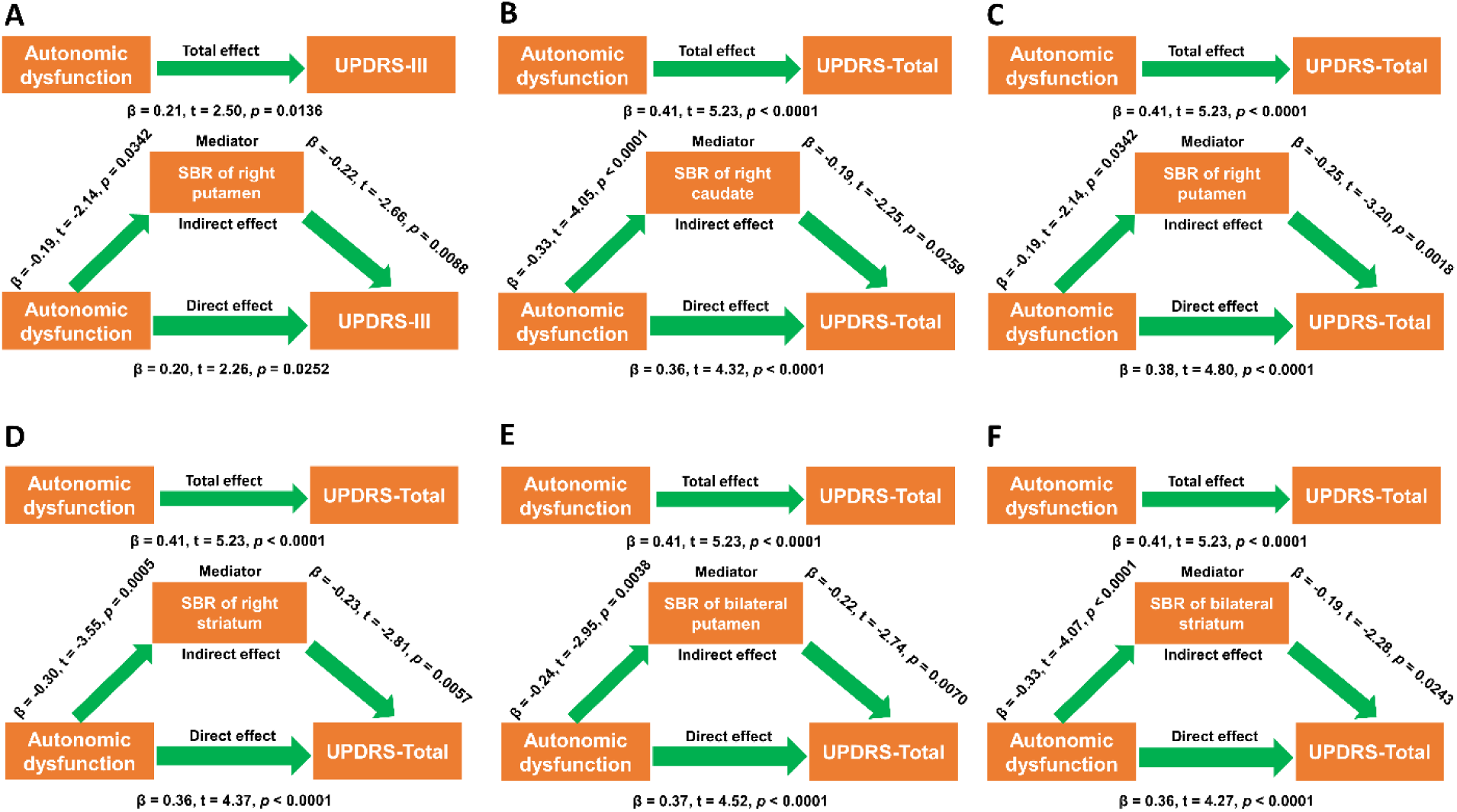
Striatal dopamine depletion mediated the effects of autonomic dysfunction on overall disease burden. (A) SBR of right putamen mediated the effects of autonomic dysfunction on UPDRS-III scores. (B-F) SBRs of right caudate (B), right putamen (C), right stratum (D), bilateral putamen (E), and bilateral stratum (F) mediated the effects of autonomic dysfunction on total UPDRS scores. During the mediation analysis, age, sex, disease duration, and years of education were included as covariates. *p* < 0.05 was considered statistically significant. Abbreviations: UPDRS, Unified Parkinson’s Disease Rating Scale; SBR, striatal binding ratio.

Age was significantly associated with both autonomic dysfunction and striatal dopamine depletion; therefore, we hypothesized that autonomic dysfunction might mediate the effects of age on striatal dopamine depletion. Actually, we found autonomic dysfunction mediated the effects of age on SBRs of bilateral caudate and striatum (Figure S3A-B).

## Discussion

In this study, we revealed that autonomic dysfunction was associated with worse motor and non-motor symptoms in PD patients. Additionally, with graphical analysis, we showed that PD patients with more severe autonomic symptoms exhibited significantly different nodal network metrics in both structural and functional network compared to patients with less autonomic symptoms. Furthermore, we demonstrated that both brain network topology and striatal dopamine depletion mediated the effects of autonomic dysfunction on clinical symptoms. Our findings provided up-to-date information to enhance the understanding of the impacts of autonomic dysfunction on clinical symptoms in PD.

### Autonomic dysfunction and clinical symptoms in PD

Autonomic dysfunction is prevalent in older adults, especially in frail elderly.^[40]^ It has been shown that age-dependent gut-to-brain or brain-to-gut propagation of α-synuclein pathology along the sympathetic and parasympathetic nervous system, leading to age-dependent dysfunction of the heart and stomach.^[11]^ In a recent study, we found age was significantly associated with SCOPA-AUT scores in PD patients,^[29]^ which was replicated in our current study showing that PD patients with worse autonomic dysfunction exhibited higher age compared to PD patients with milder autonomic symptoms. These results suggested that age was a key demographic factor significantly shaping autonomic symptoms in PD patients. We found no statistical difference in sex distribution among 3 quartile groups, which was consistent with our recent study showing that sex was not associated with autonomic symptoms in PD patients.^[29]^ In current study, we found autonomic dysfunction was positively associated with HY stages and UPDRS-III scores, which was in agreement with previous studies showing that autonomic dysfunction was associated with faster motor progression.^[9]^ We found autonomic symptoms were positively associated with ESS scores, which was supported by a recent study showing that more severe EDS in PD patients was in parallel with more worsened autonomic symptoms.^[41]^ A previous study showed that female sex, high HY stage, high UPDRS total and subtotal scores, and cognitive decline were significantly associated with depressive symptoms.^[42]^ In this study, we found autonomic symptoms were also associated with depressive symptoms, which was consistent with the results reported by a multi-center cohort study in China.^[5]^ These results indicated that autonomic dysfunction might exacerbate depressive symptoms in PD patients. We found autonomic symptoms were positively associated with RBDSQ scores, which was supported by a recent study showing that autonomic dysfunction was associated with RBDSQ scores.^[5]^ Autonomic dysfunction occurred in 83% of isolated RBD patients, and severe cardiovagal autonomic dysfunction was associated with phenoconversion to dementia with Lewy bodies but not PD,^[43]^ which indicated that autonomic dysfunction usually co-occurred with RBD in prodromal stage of PD. In this study, we found autonomic symptoms were positively associated with anxiety levels, which was in line with the results reported by previous studies.^[44, 45]^ Recently, autonomic dysfunction was found to be associated with cognitive impairment in PD patients,^[46, 47]^ which was consistent with our findings that autonomic dysfunction was negatively associated with SDMT and MoCA scores. Striatal dopamine depletion was a key pathological feature of PD. In this study, we found autonomic symptoms were negatively associated with striatum SBRs, indicating that autonomic dysfunction may be linked to dopaminergic neurodegeneration in PD patients. Indeed, a recent study has shown that autonomic symptoms were significantly correlated with lower SBRs in right caudate, which was mainly driven by gastrointestinal and cardiovascular dysfunction.^[48]^ In addition, it has also been observed that patients with pure autonomic failure exhibited similarly severe nigral and overall central dopaminergic denervation as PD patients had.^[49]^ These results suggested that autonomic dysfunction may contribute to striatal dopamine depletion in PD patients. Taken together, we concluded that autonomic dysfunction in PD was associated with more severe motor and non-motor symptoms, as well as worse dopaminergic neurodegeneration in PD patients.

### Autonomic dysfunction, structural networks, and clinical symptoms

To examine how autonomic dysfunction shaped structural and functional network metrics, we compared the group differences of structural and functional topological metrics among 3 quartile groups. For structural network, we found PD patients with more severe autonomic symptoms exhibited lower betweenness centrality and degree centrality in right superior frontal gyrus and left calcarine compared to patients with milder autonomic symptoms, which indicated that autonomic dysfunction specifically impaired the local topology of right superior frontal gyrus and left calcarine. The effect of autonomic dysfunction on local topology of right superior frontal gyrus was consistent with the lower nodal efficiency in right superior frontal gyrus. In addition, left superior frontal gyrus also showed lower degree centrality and nodal efficiency, which also supported that autonomic dysfunction may be associated with disrupted nodal efficiency of bilateral superior frontal gyri. However, using multivariate regression analysis, we found it was age but not autonomic dysfunction was significantly associated with the nodal efficiency of bilateral superior frontal gyri. In contrast, we found nodal degree centrality of left calcarine was significantly associated with SCOPA-AUT scores, which was independent of age, sex, years of education, and disease duration. In addition, we also found that degree centrality of left calcarine was negatively associated with HY stages, UPDRS-III scores, total UPDRS scores, ESS scores, and positively associated with SDMT and MoCA scores. The significant association between degree centrality of left calcarine and motor impairment was supported by a recent study showing that degree centrality at the left calcarine was negatively associated with UPDRS-III scores in PD patients without carrying A allele of *AQP4* rs162009.^[50]^ According to previous studies, the impairment of structure and function in left calcarine has been revealed in PD patients.^[51, 52]^ For instance, PD patients displayed reduced regional homogeneity in left calcarine in comparison with healthy control.^[51]^ In addition, it has also been shown that reduced gray matter volume in left calcarine was associated with poor attention and executive function, which exacerbated motor decline in PD patients.^[52]^ Therefore, it was possible that the dysfunction of left calcarine might contribute to cognitive decline, which led to impaired motor control and motor deterioration. Indeed, cognitive impairment was actually associated with motor impairment.^[53]^ In our recent study, we also observed cognitive decline in PD contributed to motor impairment in PD patients.^[54]^ According to previous studies, left calcarine was also associated with cognition decline in other diseases.^[55–57]^ For example, lower gray matter volume of left calcarine was correlated with the impairment of visuospatial ability and episodic memory in *APOE* ε4 carriers of AD.^[55]^ Additionally, it has been shown that functional connectivity of left calcarine was negatively correlated with MoCA scores in Tinnitus patients with cognitive impairment.^[56]^ Moreover, in type 2 diabetes mellitus, patients with cognitive impairment exhibited lower functional connectivity in left calcarine, which was associated with lower episodic memory performance in these patients.^[57]^. Taken together, our findings suggested that left calcarine was a key node involved in cognitive function and mediated the effects of autonomic dysfunction on motor impairment of PD patients. In addition, degree centrality of left calcarine may also be a potential imaging biomarker for disease progression in PD patients.

### Autonomic dysfunction, functional networks, and clinical symptoms

In the functional network, we found significantly different nodal network properties in multiple central autonomic regions among 3 quartile groups, such as right middle orbitofrontal gyrus, bilateral posterior cingulate cortices, bilateral rectus, and left anterior cingulate cortex. Interestingly, nodal degree centrality, nodal efficiency, and nodal local efficiency were all higher in PD patients with more severe autonomic dysfunction compared to PD patients with milder autonomic dysfunction. With association analysis, we further demonstrated that degree centrality and nodal efficiency of bilateral posterior cingulate cortices were positively associated with SCOPA-AUT scores. Therefore, bilateral posterior cingulate cortices were the major central targets modulated by autonomic dysfunction in PD patients. Using mediation analysis, we further showed that degree centrality and nodal efficiency of bilateral posterior cingulate cortices mediated the effects of autonomic dysfunction on SDMT scores, which suggested that bilateral posterior cingulate cortices contributed to cognitive decline in PD patients. These results were consistent with our recent findings that degree centrality and nodal efficiency of right posterior cingulate cortex mediated the effects of striatal dopamine depletion on SDMT scores.^[32]^ Posterior cingulate cortex was a key node in default mode network^[58]^ and essential for executive, mnemonic and spatial processing functions.^[59]^ In line with our findings, the increased function connectivity of posterior cingulate cortex in PD patients with cognitive impairment has been reported.^[60]^ Based on these results, we concluded that autonomic dysfunction negatively affected cognitive function by modulating the local topology of bilateral posterior cingulate cortices in PD patients.

### Autonomic dysfunction, striatal dopamine depletion, and clinical symptoms

As shown above, we found autonomic symptoms were negatively associated with striatal dopamine availability, indicating that autonomic dysfunction may contribute to dopaminergic neurodegeneration in PD patients.^[48]^ In a recent study, we found age was negatively associated with striatal dopamine availability,^[29]^ thus we hypothesized that autonomic dysfunction may mediated the effects of age on striatal dopamine availability. With mediation analysis, we actually demonstrated that autonomic dysfunction mediated the negative association between age and striatal dopamine availability. Indeed, it has been shown that aging promoted α-synuclein propagation from gut to brain though sympathetic and parasympathetic nerves, which resulted in peripheral autonomic dysfunction.^[11]^ According to our recent perspectives, peripheral autonomic neurodegeneration may induce α-synuclein propagation from gut to brain through autonomic nerves,^[1]^ which further triggered central dopaminergic neurodegeneration and striatal dopamine depletion.^[61, 62]^ Therefore, we concluded that autonomic dysfunction contributed to age-dependent decline of striatal dopamine availability in PD patients. In a recent study, we showed that striatal dopamine availability was associated with both UPDRS-III and total UPDRS scores of PD patients.^[32]^ In this study, we revealed autonomic dysfunction was associated motor and non-motor symptoms, as well as striatal dopamine availability of PD patients. Therefore, we hypothesized that striatal dopamine depletion may mediate the effects of autonomic dysfunction on motor symptoms and overall disease burden. Actually, we found SBR of right caudate specifically mediated the effects of autonomic dysfunction on UPDRS-III scores. In addition, we also demonstrated that SBRs of right caudate, right putamen, right striatum, bilateral putamen, and bilateral striatum mediated the effects of autonomic dysfunction on total UPDRS. Taken together, our study suggested that autonomic dysfunction exacerbated disease progression by triggering striatal dopamine depletion in PD patients.

### Strengths and limitations of the study

In this study, we found more severe autonomic symptoms were associated with worsened motor and non-motor symptoms, as well as striatal dopamine depletion in PD patients, which suggested that autonomic dysfunction was a potential predictor for disease progression in PD patients. Using graphical analysis, we revealed that autonomic dysfunction shaped both structural and functional network topology, implying that autonomic dysfunction played a special role in the heterogeneity of brain network topology in PD patients. Specifically, we demonstrated that both structural and functional network metrics mediated the effects of autonomic dysfunction on clinical symptoms of PD patients, which provided network-level mechanisms to explain the effects of autonomic dysfunction on clinical manifestations of PD patients. Finally, we revealed that age-dependent autonomic dysfunction contributed striatal dopamine depletion in PD patients, which further exacerbated motor and cognitive impairments in PD patients. Taken together, our current study suggested that both brain network topology and striatal dopamine depletion contributed to motor and non-motor deterioration associated with autonomic dysfunction in PD patients. Because our findings were mainly based on cross-section analysis, our results were required to be confirmed in future longitudinal studies.

### Conclusions

PD patients with worse autonomic symptoms exhibited more severe motor and non-motor manifestations. In addition, autonomic dysfunction significantly shaped structural and functional network topology in PD patients. Differential structural and functional network properties were associated with cognitive and motor decline of PD patients.

## Acknowledgments

Data used in the preparation of this article were obtained from the Parkinson’s Progression Markers Initiative (PPMI) database (www.ppmiinfo.org/data). We thank the share of PPMI data by all the PPMI study investigators. PPMI – a public-private partnership – is funded by the Michael J. Fox Foundation for Parkinson’s Research and funding partners, which can be found at www.ppmiinfo.org/fundingpartners.

## Data Availability Statement

All the raw data used in the preparation of this Article were downloaded from PPMI database (www.ppmi-info.org/data). All data produced in the present study are available upon reasonable request to the authors.

## Supporting Information

Additional supporting information may be found online in the Supporting Information section at the end of the article.

