## Supplementary materials for "Both brain network topology and striatal dopamine depletion mediate the effects of autonomic dysfunction on disease burden of Parkinson’s disease"

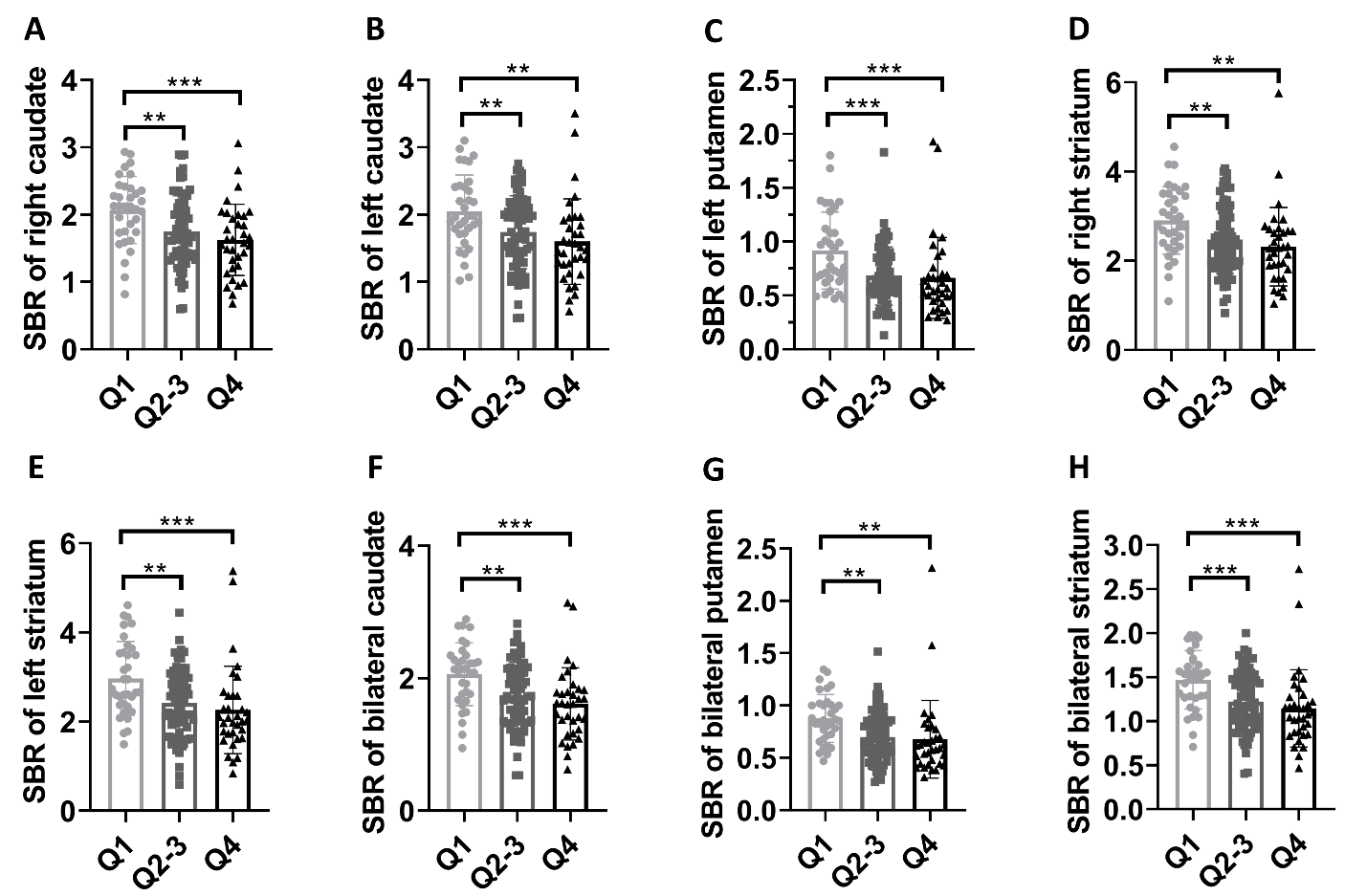

**Supplementary Figure 1: Group differences in stratum SBRs.** (A-H) Group differences in SBRs of right caudate (A), left caudate (B), left putamen (C), right stratum (D), left stratum (E), bilateral caudate (F), bilateral putamen (G), and bilateral stratum (H). One-way ANOVA followed by Tukey’s post hoc test (Q1 group *vs* Q2-3 group *vs* Q4 group) were conducted to compare clinical variables. *p* < 0.05 was considered statistically significant. ***p* < 0.01, ****p* < 0.001. Abbreviations: SBR, striatal binding ratio.

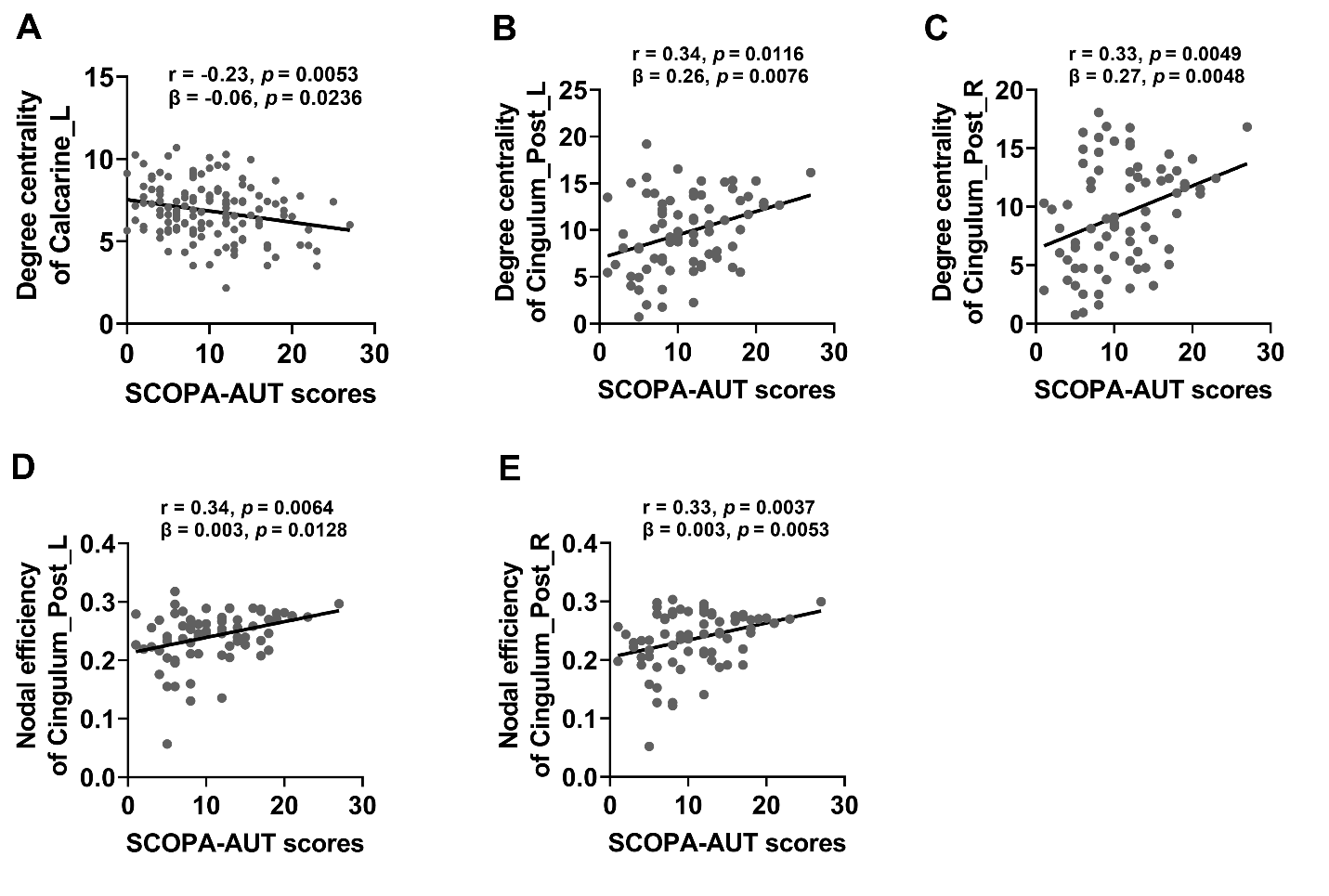

**Supplementary Figure 2: Associations between nodal network metrics and SCOPA-AUT scores.** (A) SCOPA-AUT scores were negatively associated with degree centrality of left calcarine in structural network. (B-C) SCOPA-AUT scores were positively associated with degree centrality of left posterior cingulate cortex (B) and right posterior cingulate cortex (C). (D-E) SCOPA-AUT scores were positively associated with nodal efficiency of left posterior cingulate cortex (D) and right posterior cingulate cortex (E). The association analysis between graphical network metrics and clinical assessments was conducted by Pearson correlation method and multivariate regression analysis with age, sex, disease duration, and years of education as covariates. FDR-corrected *p* < 0.05 was considered statistically significant. Abbreviations: SCOPA-AUT, Scale for Outcomes in Parkinson's Disease-Autonomic.

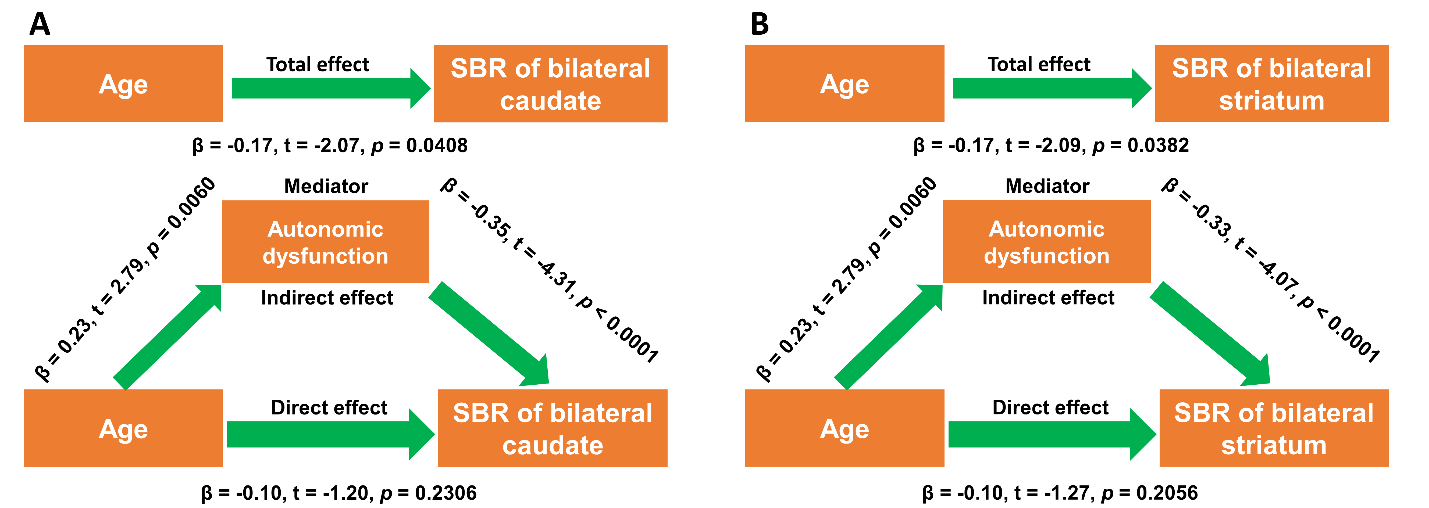

**Supplementary Figure 3: Autonomic dysfunction mediated the effects of age on striatal dopamine depletion.** (A-B) Autonomic dysfunction mediated the effects of age on SBRs of bilateral caudate and striatum. During the mediation analysis, age, sex, disease duration, and years of education were included as covariates. *p* < 0.05 was considered statistically significant. Abbreviations: SBR, striatal binding ratio.
